# Non-contact and non-constraining monitoring of the respiratory rate including sleep disordered breathing using ultra-wideband radar

**DOI:** 10.1101/2024.07.08.24310110

**Authors:** Chin Kazuo, Okumura Shigeaki, Endo Daisuke, Nagata Kazuma, Ito Tatsuya, Murase Kimihiko, Sunadome Hironobu, Hoshi Mamiko, Hiranuma Hisato, Kozu Yutaka, Sato Susumu, Hirai Toyohiro, Gon Yasuhiro, Sakamoto Takuya, Taki Hirofumi, Akahoshi Toshiki

**Author notes:** Corresponding author Kazuo Chin Department of Sleep Medicine and Respiratory Care, Division of Respiratory Medicine, Nihon University of Medicine. 30-1 Ohyaguchi kami-cho, Itabashi-ku, Tokyo 173-8610, Japan (EXT 2401 · 2402).

## Abstract

**Background:** The respiratory rate (RR) is a critical vital sign controlled by not only metabolic factors but behavior while awake. The prevalence of obstructive sleep apnea (OSA) is remarkably high. Therefore, a non-constraining and non-contact respiratory monitor to measure the RR both while awake and asleep is preferable.

**Methods:** A millimeter wave radar (MWR) device was used for RR monitoring of participants suspected of OSA while awake (supine before and after polysomnography [PSG], sitting, and positioned on both sides) and asleep. Apnea and hypopnea during 1 hour of sleep (AHI) by PSG was compared with 1 hour of respiratory events (REI) judged by MWR. Portable monitoring (PM) and percutaneous arterial O_2_ (SpO_2_) monitoring were done simultaneously.

**Results:** Correlations between the RR by MWR and respiratory inductance plethysmography at PSG while awake in every measured position including the supine position before and after PSG were very high (r=0.92∼0.99) (n=60). The REI by MWR were significantly correlated with AHI determined by PSG, PM, or SpO_2_ monitoring (p<0.001). Brand-Altman plot showed that the MWR for AHI monitoring was acceptable. Predicted AHI by MWR relative to PSG was almost the same as with PM or SpO_2_ monitoring.

**Conclusions:** The developed MWR respiratory monitor was useful during wakefulness and sleep, including detection of apnea and hypopnea. This system can be useful in multiple medical settings such as critical care with and without sleep apnea, pandemic infection, elder care at home, etc. Trial registration number: UMIN000045833 (http://www.umin/ac.jp)

## Introduction

Respiration is controlled by not only chemical or metabolic factors but nonchemical means including behavior while awake.^1–3^ To measure the respiratory rate (RR) during awake periods, a non-constraining and non-contact respiratory monitor is desired. In measuring the RR during sleep, it is important to detect sleep disordered breathing (SDB), mainly obstructive sleep apnea (OSA) and hypopnea^4^, as an extremely high prevalence of OSA was shown.^5–7^ A non-contact respiratory monitoring device to measure RR during wakefulness and sleep would be extremely useful in hospital settings including those related to critical care, post-operation, infectious diseases such as COVID-19, injuries, or sleep apnea diagnosis and at home such as with elder care.

There are 2 types of non-contact monitoring such as radar and a camera.^8,9^ For privacy and information obtained through imaging, radar seems preferable. Radar sends out electromagnetic signals indicating changes in ribcage and abdominal displacements due to respiration. RR can be measured by the millimeter wave radar (MWR) system. We recently reported that this system could detect sleep apnea.^10,11^ A comprehensive review of doppler radar-based non-contact monitoring for sleep apnea was also published.^12^ Recently the sleep state of healthy individuals or RR during polysomnography (PSG) in patients with OSA by radar was reported.^13–15^ However, RR measurements during the awake state and sleep apnea simultaneously in clinical settings with PSG and portable monitors (PM) used to diagnose OSA were not reported. We hypothesized that non-constraining and non-contact respiratory monitoring based on an MWR system can detect not only RR without sleep apnea and hypopnea (SAH) and SAH during sleep in clinical settings as well as PM.^4^ To test this hypothesis, we monitored participants in our sleep clinic suspected of having OSA.

## Methods

### Participants and Study Design

Study participants were consecutive patients suspected as having SAH and scheduled for MWR monitoring during polysomnography (PSG). After obtaining approval, we set up the PSG monitor and radar device (Figure 1-a). Before PSG, we monitored breathing for 3 min while the participant was sitting, supine, and lying on both sides (Figure 1-b). Following these RR measurements, we performed PSG as usual. The RR was measured again in the supine position after morning PSG and also in some participants in the supine position with or without a blanket before sleep (Figures 1-a, S2). This study was approved by the Nihon University Itabashi Hospital Clinical Research Review Board (RK-210914-5) and was registered with the University Hospital Medical Information Network (in Japan) Clinical Trials Registry (identification number UMIN000045833). All participants provided written informed consent.

**Figure 1.**
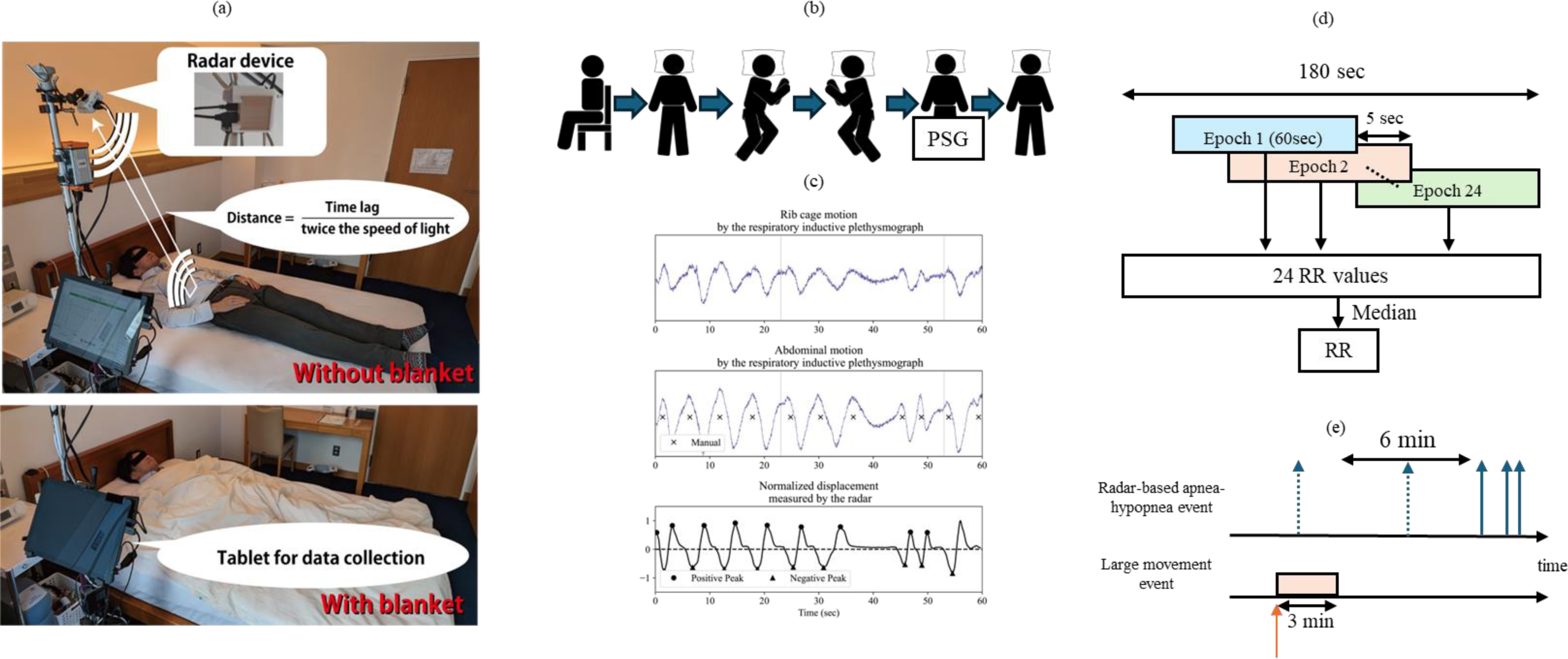
a) The millimeter wave radar (MWR) near the bed The MWR was placed near the bed attended by one of the authors. He was with or without a blanket. The radar device and tablet for data collection are shown. The distance between the radar and chest wall was calculated as distance=(Time lag)/(twice the speed of light). b) Positions for measurements. c) Ribcage and abdominal motions by respiratory inductive plethysmography (RIP) and normalized displacement by radar A technician recognized x as a respiration by abdominal or ribcage RIP. Positive (●) and negative (▲) peaks by the radar are shown. d) Method of calculation of the respiration rate (RR) by radar. After 5 s from the start of the radar system, a positive or negative peak was automatically detected at a minimal interval of 1.5 s. Each positive or negative median value during 60 s was calculated and the number of respirations in which the length from positive to negative peak was less than 10% of each min median value was eliminated. The number of residual positive or negative peaks was considered to be the RR. The radar system calculated the median RR every 5 s using last 60 s data following the first 60 s. Total duration of the measurement was 180 s. e) Method of calculations of apnea and hypopnea episodes. The algorithm mistakenly detects apnea and hypopnea when large body posture changes occur during sleep such as position changes and large movements following prolonged apnea or severe hypoxemia. Therefore, if a large displacement occurred, the apnea and hypopnea events (dotted arrows) were discarded for the 3 min following the last large movement (red arrow). In addition, one solitary apnea or hypopnea event that occurred every 6 min was discarded. Horizonal arrows show the counted apnea or hypopnea episodes. PSG: polysomnography, RR: respiratory rate

## Collection of Data

### Polysomnography, portable monitoring, and continuous pulse-oximetry monitoring Polysomnography

PSG (Alice 6; Philips Respironics, Murrysville, PA, USA) was performed from 2200 until 0600 h using standard techniques. Airflow was monitored with a nasal pressure transducer and an oronasal thermal sensor. Ventilation was monitored with respiratory inductive plethysmography (RIP) (Pro-Tech Services Inc, Murrysville, PA, USA). Apnea was defined as a ≥90% reduction in airflow for more than 10 s, and hypopnea was defined as a ≥30% reduction in airflow for >10 s accompanied by ≥3% oxygen desaturation or arousal.^16^ The apnea and hypopnea index (AHI) indicates the number of apnea and hypopnea episodes per 1 h of sleep.

### Portable-monitoring recording and continuous pulse-oximetry monitoring

We used the Pulsleep LS-140 (Fukuda Denshi Co., Ltd., Tokyo, Japan) as the PM.^17^ This is a type 3 monitor released in 2017. This device was run with PSG concurrently. Respiratory events per 1 h of measurement were defined as the respiratory event index (REI). REI from LS-140 was checked by a trained technician. From the pulse-oximetry (SpO_2_) monitor attached to the LS-140, we calculated the 3% oxygen desaturation index (3%ODI) separately.

## Data Collection by Radar

### Measurement of the respiratory rate by radar while awake

The radar (Figure 1-a: T14_01120112_2D, S-Takaya Electronics Industry Company, Ltd., Okayama, Japan) used was reported to present no health hazards to humans.^18^ Ribcage and abdominal movements by RIP during PSG monitoring and amplitude changes according to the radar are shown in Figure 1-c. The RR by IP was calculated by a technician. We measured the RR by radar as follows: 1) 5 s after the radar system started, a positive or negative peak was automatically detected at a minimal interval of 1.5 s, making it possible to measure RRs up to 40 bpm with this system; 2) each positive or negative median value during 60 s was calculated and the number of respirations of which the length from the positive to negative peak was cut to <10% of each min median value recorded; 3) the residual positive or negative peak was considered to be the RR (Figure 1-c); and 4) the radar system calculated the RR every 5 s using the last 60 s data following the first 60 s. Total duration of the measurement was 180 s. The PSG technicians checked the RR (x) by ribcage or abdominal RIP (31.3% by ribcage and 68.7% by abdominal RIP, n=300, Figure 2). We finally compared the median RRs obtained manually with those from radar (Figure 1-c). Following RR measurements in the supine position before PSG, RRs from 15 of the 60 participants were measured with a blanket (Figures 1-a and S2). After 5 s from the re-start of the radar system, we measured the RR with the blanket for almost 3 min the same way as without the blanket.

**Figure 2.**
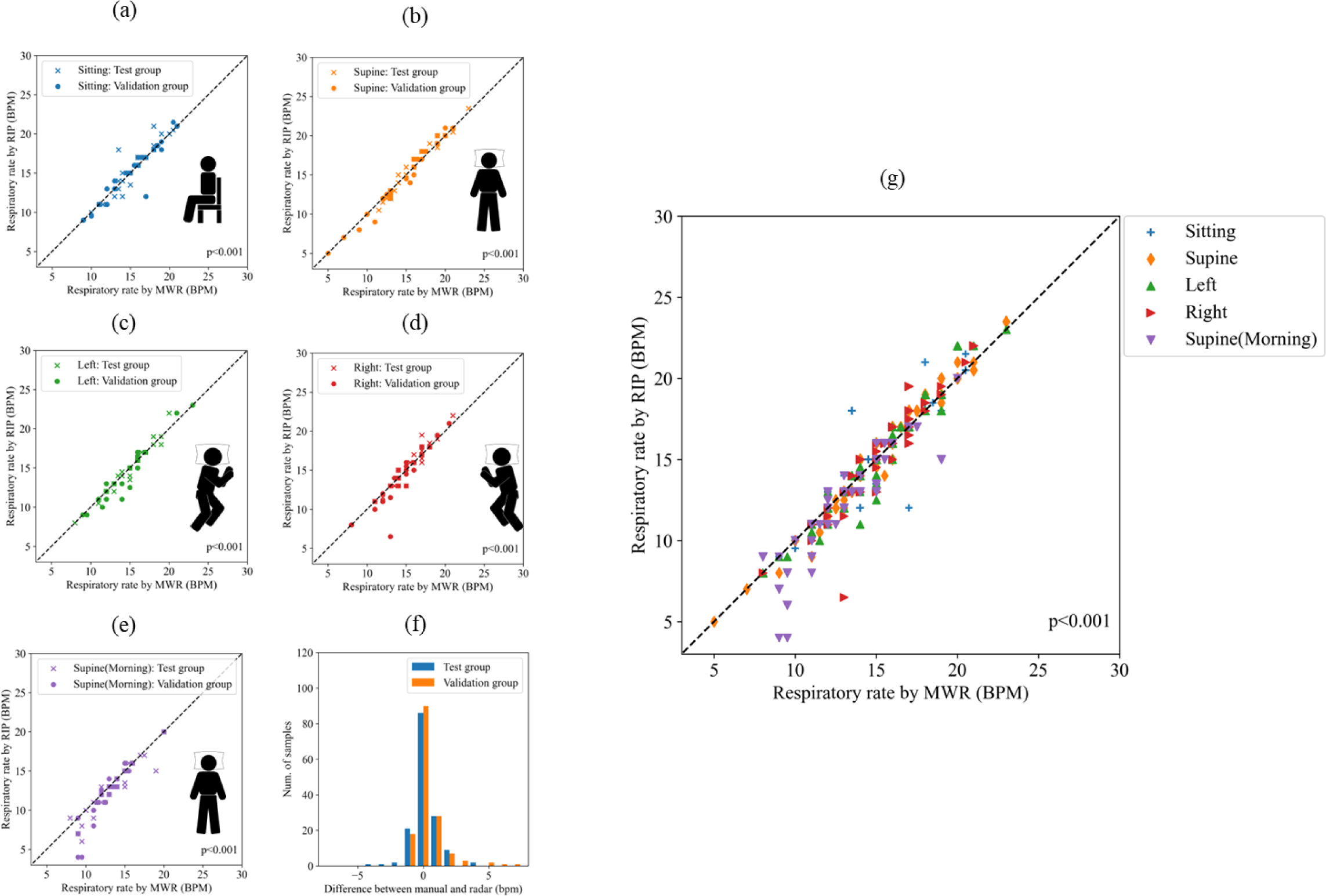
Respiration rates (RR) by radar (X axis) and manually calculated from respiratory inductance plethysmography (RIP) by a technician (Y axis) Participants’ RR (Test group: first 30 participants, Validation group: next 30 participants) were calculated while sitting (a), supine before (b) and after polysomnography (e), right and left lateral (d, e) positions and including all positions (g). Differences in RR between manual and radar measurements (X axis) and the number of participants (Y axis) are shown (f). Almost all the differences between the radar and manual measurements were within 2 and mostly 1 times/min (f). RIP; respiratory inductance plethysmograph, RR, respiratory rate; MWR, millimeter wave radar, BPM: breaths per minute.

### Methods of measurements of sleep disordered breathing (apnea and hypopnea) by radar

The radar-based method for detecting sleep disordered breathing (SDB) was explained previously.^10,11^ In the present study, respiratory events were detected also based on the Expectation-Maximization (EM) Algorithm.^11^ Briefly, the EM algorithm estimates parameters for stochastic models of respiratory movements. It can distinguish between normal breathing and apnea or hypopnea and detect apnea and hypopnea without using many datasets of non-participants for training or threshold values. When we assume that the amplitudes of body surface movements caused by respiratory effort show a Gaussian distribution, amplitudes that include both apnea or hypopnea and normal breathing can be represented as a Gaussian mixture model. SDB can be detected based on the difference in two Gaussian distributions estimated by the EM algorithm. The threshold for the ratio of two Gaussian distributions in the EM algorithm to detect SDB events was set at 0.7 (Figure S1). We used 60 s of radar data for radar-based apnea and hypopnea event detection and the analysis epoch was overlapped every 5 s. As mentioned above, the algorithm detects changes in amplitude as a radar-based apnea and hypopnea event. Therefore, the algorithm mistakenly detects apnea and hypopnea when large body posture changes occur during sleep such as large movements with arousals or position changes following prolonged apnea or severe hypoxemia. We could check large movements by arousals or position changes by two types of sample duration (0.32 or 1.28 s) by the 100 Hz radar. For the first 30 participants, we applied the stabilization process (Figure 1-e). We eliminated data following certain big movements because subsequent respiration amplitudes were very unstable. We explored the duration that is most suitable to those data to approximate the AHI by PSG. We found that 3 min was the most suitable to match REI by radar to the AHI by PSG. In addition, one apnea or hypopnea episode that occurred once every 6 min was not calculated (Figure 1-e). Radar started and stopped when the PSG started and stopped. In summary, there were 4 parameters by the radar as follows:

[1] :total number of reliable detected events
[2] : total min of radar measurement
[3] : summation of each duration of large movement+3 (min)
[4] : total number of solitary apnea or hypopnea events during 6 min

The REI according to the radar was finally defined by ([1]-[4])/([2]-[3])*60

### Statistical analysis

Continuous variables are expressed as means ± standard deviation (SD) and categorical variables as counts and percentages. Differences between AHI and three measurements (REI by radar, PM, 3% ODI) are shown as mean differences ± SD. Two-group comparisons used the unpaired Student’s t-test, and categorical data were analyzed with the Chi-square or Fisher’s exact test. Pearson correlation coefficients assessed correlations between RR determined by radar and manual data obtained by trained technicians and AHI correlations with the three measurements. These three measurements were compared using ANOVA. A P value <0.05 indicated statistical significance. Bland-Altman plots for method agreement were generated using Python 3.8.16 with NumPy 1.24.2 and Matplotlib 3.4.3.^19,20^ Other statistical analyses were done using EZR on R Commander 2.8-0 and R 4.2.2.^21^

### Sample size determination

If there is no difference among the PM, ODI, and radar measurements for AHI from 0 to 60, then 22 (standard deviation, SD=5) or 32 (SD=6) paired participants are required to be 90% certain that the limits of a two-sided 90% confidence interval will exclude a difference in means >5. We preferred SD=6.

## Results

### Participants’ characteristics

Participants were 89 persons suspected to have sleep apnea. We analyzed participants whose AHI ranged from 0 to 60. In 22 participants the AHI was **>**60. In the remaining 67 participants, the first 30 patients were randomly selected as a “test group” with an almost equal distribution and included those with normal 0≤AHI<5, mild 5≤ AHI<15, moderate 15≤AHI<30, and severe AHI≥30 among 7, 8, 6, and 9 persons, respectively. The last 30 participants with normal, mild, moderate, and severe values in 8, 9, 6, and 7 participants, respectively, became the “validation group”. Finally, 7 participants were excluded from the analysis whose AHI was 4.7, 6.4, 8, 9.4, 16.9, 24 and 30.5, respectively (Table 1). From test group data, we calculated apnea and hypopnea values by radar as described above and validated the data in the final 30 patients. Characteristics of the study participants are summarized in Table 1.

**Table 1.**
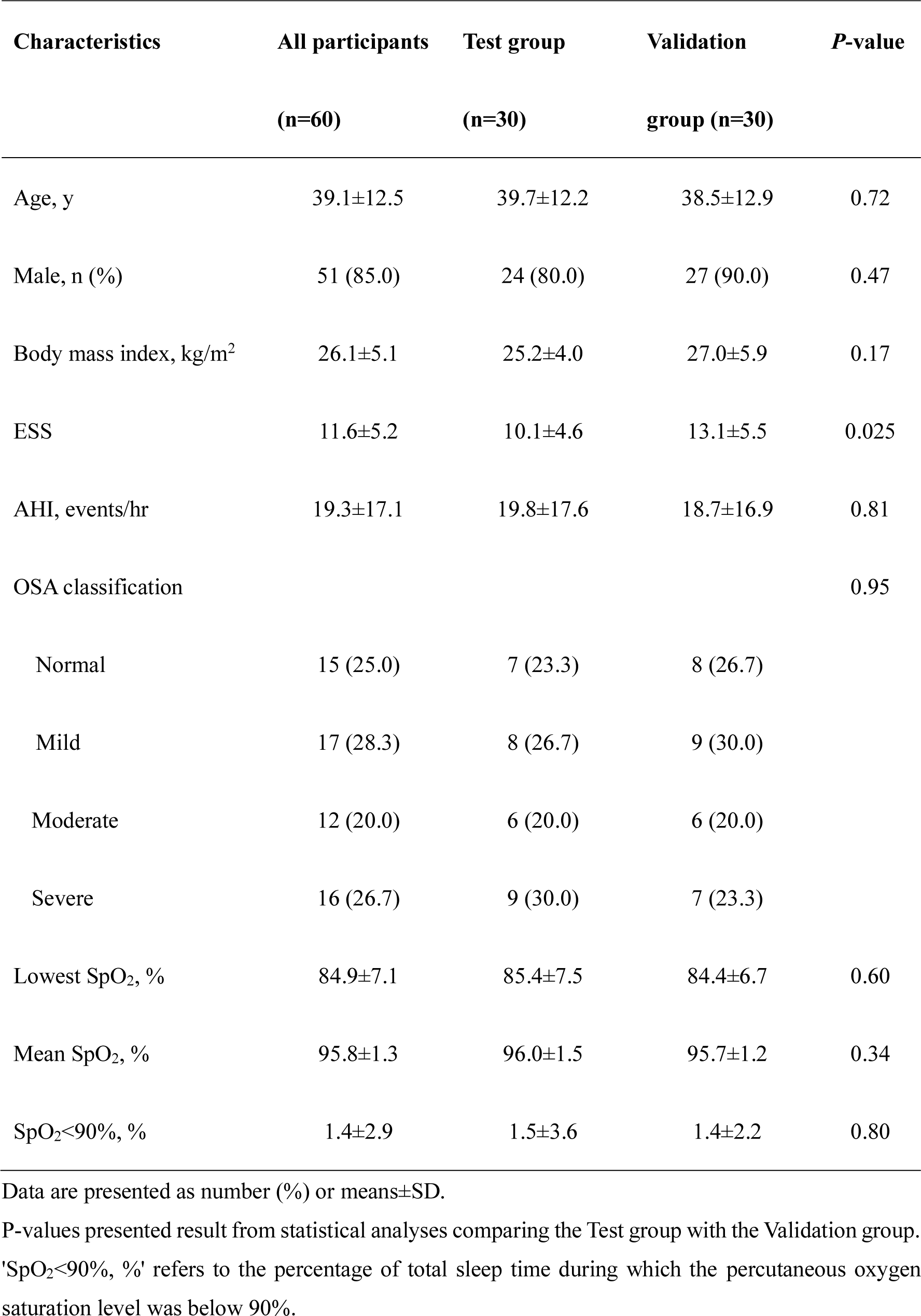
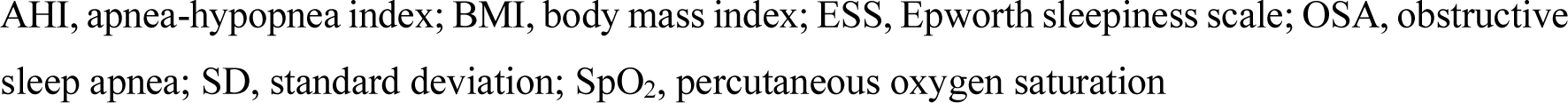
Participant characteristics between test and validation groups.

| Characteristics | All participants<br>(n=60) | Test group<br>(n=30) | Validation<br>group (n=30) | *P*-value |
| --- | --- | --- | --- | --- |
| Age, y | 39.1±12.5 | 39.7±12.2 | 38.5±12.9 | 0.72 |
| Male, n (%) | 51 (85.0) | 24 (80.0) | 27 (90.0) | 0.47 |
| Body mass index, kg/m<sup>2</sup> | 26.1±5.1 | 25.2±4.0 | 27.0±5.9 | 0.17 |
| ESS | 11.6±5.2 | 10.1±4.6 | 13.1±5.5 | 0.025 |
| AHI, events/hr | 19.3±17.1 | 19.8±17.6 | 18.7±16.9 | 0.81 |
| OSA classification |  |  |  | 0.95 |
| Normal | 15 (25.0) | 7 (23.3) | 8 (26.7) |  |
| Mild | 17 (28.3) | 8 (26.7) | 9 (30.0) |  |
| Moderate | 12 (20.0) | 6 (20.0) | 6 (20.0) |  |
| Severe | 16 (26.7) | 9 (30.0) | 7 (23.3) |  |
| Lowest SpO<sub>2</sub>, % | 84.9±7.1 | 85.4±7.5 | 84.4±6.7 | 0.60 |
| Mean SpO<sub>2</sub>, % | 95.8±1.3 | 96.0±1.5 | 95.7±1.2 | 0.34 |
| SpO<sub>2</sub><90%, % | 1.4±2.9 | 1.5±3.6 | 1.4±2.2 | 0.80 |

Data are presented as number (%) or means±SD.
P-values presented result from statistical analyses comparing the Test group with the Validation group. 'SpO<sub>2</sub><90%, %' refers to the percentage of total sleep time during which the percutaneous oxygen saturation level was below 90%.
AHI, apnea-hypopnea index; BMI, body mass index; ESS, Epworth sleepiness scale; OSA, obstructive sleep apnea; SD, standard deviation; SpO<sub>2</sub>, percutaneous oxygen saturation

### Respiratory rate monitoring with participant awake in each position (Figure 2 and Table 2-a)

First, we measured the RR with participants in the supine position with or without a blanket. We ascertained that the radar could detect the RR accurately despite the blanket (n=15) (without blanket r=0.98, with blanket =0.96, p<0.001) (Figure S2). Figure 2 and Table 2-a show the RR of participants determined by radar or RIP at each position (test group (X): n=30 and validation group (●): n=30). Correlations were significantly high (p<0.001). The correlations between the AHI and REI by radar, PM, or 3%ODI in test groups (n=30), validation groups (n=30), and the total participant group (n=60) were also highly significant p<0.001 (Table 2-b and Figure S3). In addition, one way analysis of variance showed that differences in REI among radar, PM, and 3%ODI were insignificant (Table 2-b). Figure 3 shows the Bland-Altman plot between the AHI (AHI=0∼60 or 0∼30) and REI by radar, PM, or 3% ODI. Visual inspection of the Bland-Altman plot revealed a low percentage of data points outside of the 95%CI in the radar measurements. One data in Figures 3-e and f was out of range.

**Figure 3.**
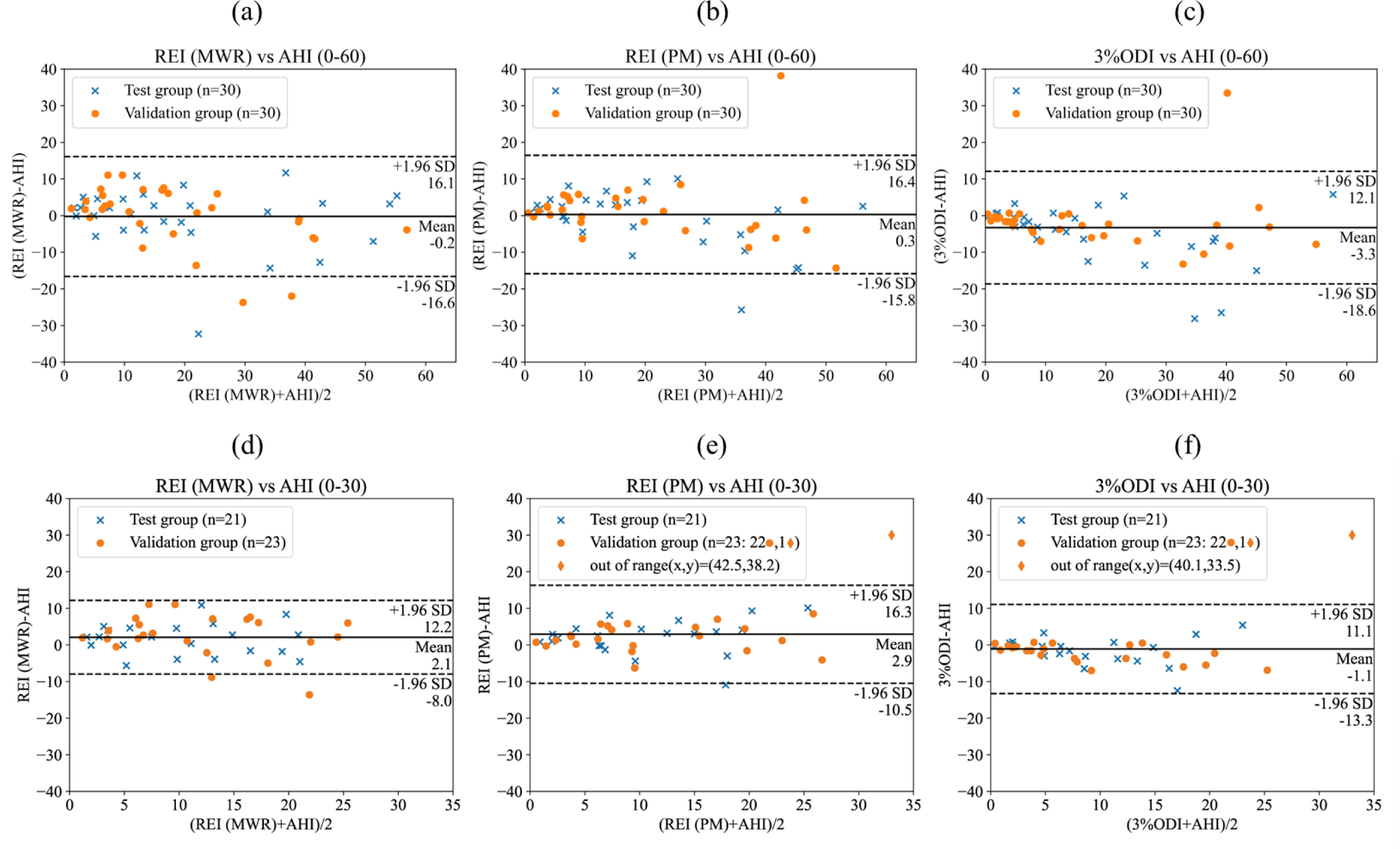
Bland-Altman plot between apnea hypopnea index (AHI=0∼60 or 0∼30) and respiratory event index (REI) by radar, portable monitor (PM) or 3% oxygen desaturation index (ODI). The differences between AHI and REI by radar, PM or 3% ODI were plotted against the mean of the 2 measurements. Solid horizontal line indicates the mean value of (REI by radar-AHI), (REI by PM-AHI) or (3%ODI – AHI), and the 2 dotted lines indicate the 95% confidence interval (mean difference ± 1.96 SD). Figures (e) and (f) show that one respiration was in out of range. AHI: apnea hypopnea index, MWR: millimeter wave radar.

**Table 2.**
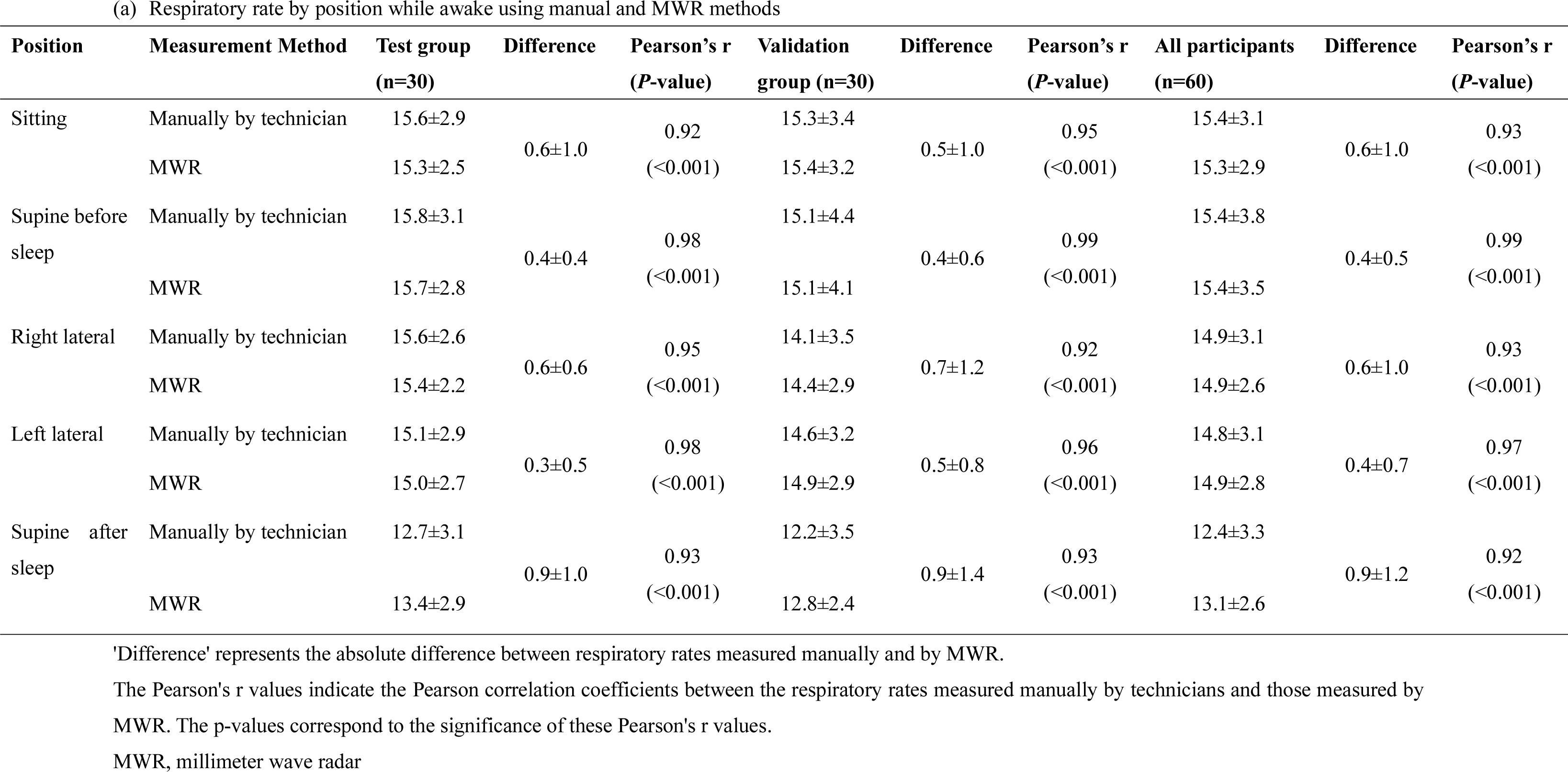
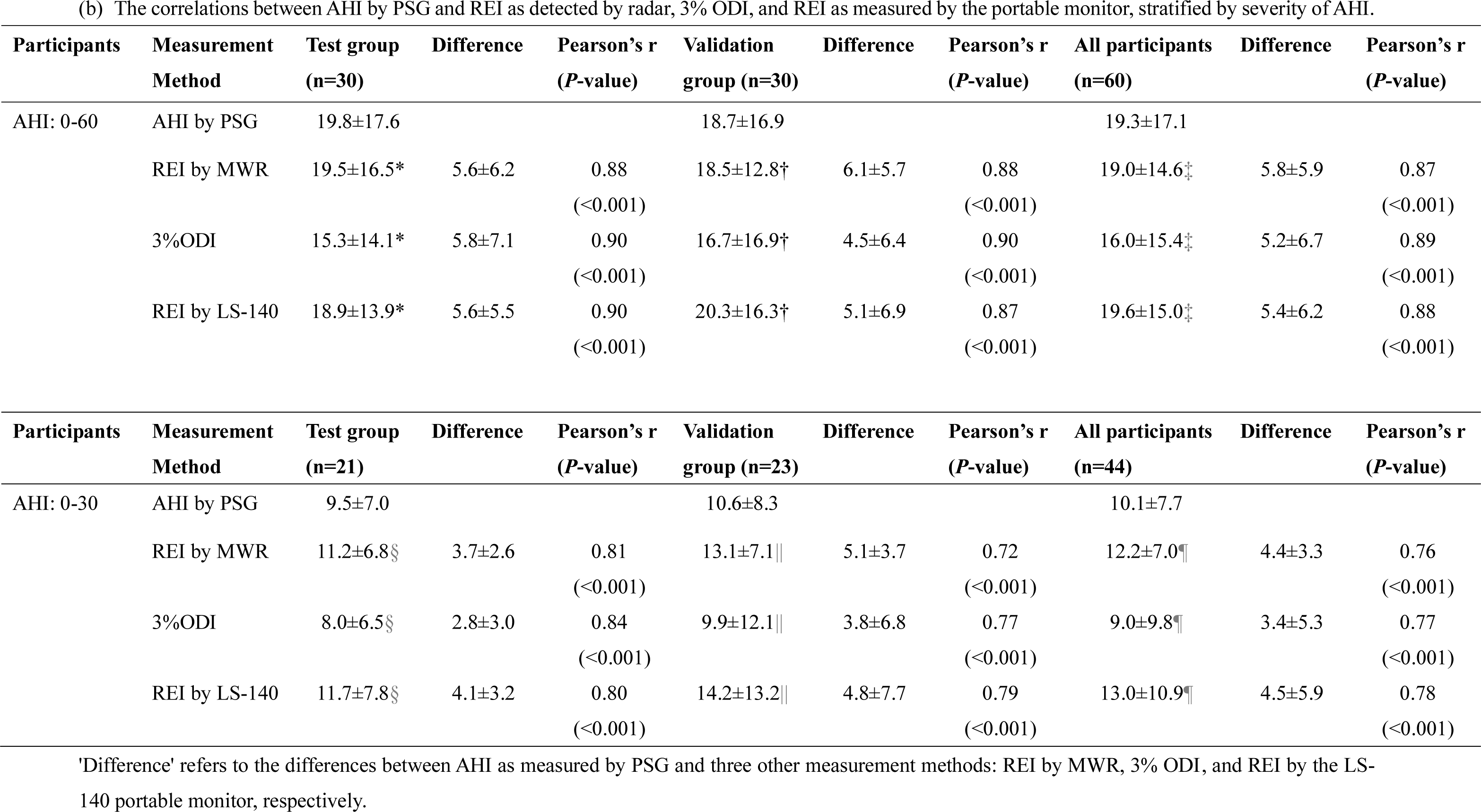
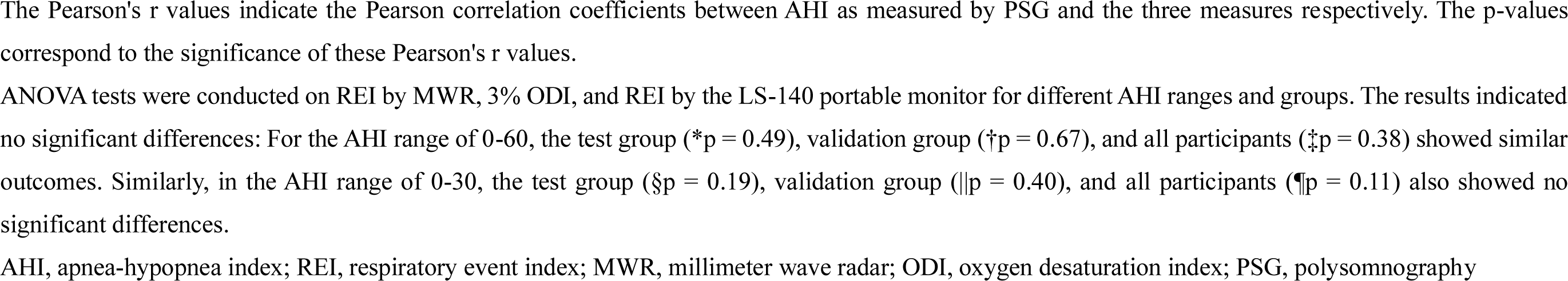
(a) Respiratory rate by position while awake using manual and MWR methods.

| Position | Measurement Method | Test group<br>(n=30) | Difference | Pearson's r<br>(*P*-value) | Validation<br>group (n=30) | Difference | Pearson's r<br>(*P*-value) | All participants<br>(n=60) | Difference | Pearson's r<br>(*P*-value) |
| --- | --- | --- | --- | --- | --- | --- | --- | --- | --- | --- |
| Sitting | Manually by technician | 15.6±2.9 | 0.6±1.0 | 0.92<br>(<0.001) | 15.3±3.4 | 0.5±1.0 | 0.95<br>(<0.001) | 15.4±3.1 | 0.6±1.0 | 0.93<br>(<0.001) |
|  | MWR | 15.3±2.5 |  |  | 15.4±3.2 |  |  | 15.3±2.9 |  |  |
| Supine before<br>sleep | Manually by technician | 15.8±3.1 | 0.4±0.4 | 0.98<br>(<0.001) | 15.1±4.4 | 0.4±0.6 | 0.99<br>(<0.001) | 15.4±3.8 | 0.4±0.5 | 0.99<br>(<0.001) |
|  | MWR | 15.7±2.8 |  |  | 15.1±4.1 |  |  | 15.4±3.5 |  |  |
| Right lateral | Manually by technician | 15.6±2.6 | 0.6±0.6 | 0.95<br>(<0.001) | 14.1±3.5 | 0.7±1.2 | 0.92<br>(<0.001) | 14.9±3.1 | 0.6±1.0 | 0.93<br>(<0.001) |
|  | MWR | 15.4±2.2 |  |  | 14.4±2.9 |  |  | 14.9±2.6 |  |  |
| Left lateral | Manually by technician | 15.1±2.9 | 0.3±0.5 | 0.98<br>(<0.001) | 14.6±3.2 | 0.5±0.8 | 0.96<br>(<0.001) | 14.8±3.1 | 0.4±0.7 | 0.97<br>(<0.001) |
|  | MWR | 15.0±2.7 |  |  | 14.9±2.9 |  |  | 14.9±2.8 |  |  |
| Supine after<br>sleep | Manually by technician | 12.7±3.1 | 0.9±1.0 | 0.93<br>(<0.001) | 12.2±3.5 | 0.9±1.4 | 0.93<br>(<0.001) | 12.4±3.3 | 0.9±1.2 | 0.92<br>(<0.001) |
|  | MWR | 13.4±2.9 |  |  | 12.8±2.4 |  |  | 13.1±2.6 |  |  |

| Participants | Measurement Method | Test group (n=30) | Difference | Pearson's r (P-value) | Validation group (n=30) | Difference | Pearson's r (P-value) | All participants (n=60) | Difference | Pearson's r (P-value) |
| --- | --- | --- | --- | --- | --- | --- | --- | --- | --- | --- |
| AHI: 0-60 | AHI by PSG | 19.8±17.6 |  |  | 18.7±16.9 |  |  | 19.3±17.1 |  |  |
| | REI by MWR | 19.5±16.5* | 5.6±6.2 | 0.88<br>($<0.001$) | 18.5±12.8† | 6.1±5.7 | 0.88<br>($<0.001$) | 19.0±14.6‡ | 5.8±5.9 | 0.87<br>($<0.001$) |
| | 3%ODI | 15.3±14.1* | 5.8±7.1 | 0.90<br>($<0.001$) | 16.7±16.9† | 4.5±6.4 | 0.90<br>($<0.001$) | 16.0±15.4‡ | 5.2±6.7 | 0.89<br>($<0.001$) |
| | REI by LS-140 | 18.9±13.9* | 5.6±5.5 | 0.90<br>($<0.001$) | 20.3±16.3† | 5.1±6.9 | 0.87<br>($<0.001$) | 19.6±15.0‡ | 5.4±6.2 | 0.88<br>($<0.001$) |
| Participants | Measurement Method | Test group (n=21) | Difference | Pearson's r (P-value) | Validation group (n=23) | Difference | Pearson's r (P-value) | All participants (n=44) | Difference | Pearson's r (P-value) |
| AHI: 0-30 | AHI by PSG | 9.5±7.0 |  |  | 10.6±8.3 |  |  | 10.1±7.7 |  |  |
| | REI by MWR | 11.2±6.8§ | 3.7±2.6 | 0.81<br>($<0.001$) | 13.1±7.1\|\| | 5.1±3.7 | 0.72<br>($<0.001$) | 12.2±7.0¶ | 4.4±3.3 | 0.76<br>($<0.001$) |
| | 3%ODI | 8.0±6.5§ | 2.8±3.0 | 0.84<br>($<0.001$) | 9.9±12.1\|\| | 3.8±6.8 | 0.77<br>($<0.001$) | 9.0±9.8¶ | 3.4±5.3 | 0.77<br>($<0.001$) |
| | REI by LS-140 | 11.7±7.8§ | 4.1±3.2 | 0.80<br>($<0.001$) | 14.2±13.2\|\| | 4.8±7.7 | 0.79<br>($<0.001$) | 13.0±10.9¶ | 4.5±5.9 | 0.78<br>($<0.001$) |

'Difference' refers to the differences between AHI as measured by PSG and three other measurement methods: REI by MWR, 3% ODI, and REI by the LS-140 portable monitor, respectively.
The Pearson's r values indicate the Pearson correlation coefficients between AHI as measured by PSG and the three measures respectively. The p-values correspond to the significance of these Pearson's r values.
ANOVA tests were conducted on REI by MWR, 3% ODI, and REI by the LS-140 portable monitor for different AHI ranges and groups. The results indicated no significant differences: For the AHI range of 0-60, the test group (\*p = 0.49), validation group (†p = 0.67), and all participants (‡p = 0.38) showed similar outcomes. Similarly, in the AHI range of 0-30, the test group (§p = 0.19), validation group (||p = 0.40), and all participants (¶p = 0.11) also showed no significant differences.
AHI, apnea-hypopnea index; REI, respiratory event index; MWR, millimeter wave radar; ODI, oxygen desaturation index; PSG, polysomnography

## Discussion

We developed a non-contact and non-constraining monitoring device using MWR to assess the RR, including sleep apnea. It can accurately measure the RR at any position, including sitting, supine, and lateral positions with or without a blanket (Table 2-a and Figures 2 and S2). It also detected SDB, mainly sleep apnea and hypopnea, as accurately as a PM or SpO_2_ monitoring (Table 2-b, and Figure S3) at the simultaneous PSG setting. Bland-Altman analysis indicated the same results when the AHI was from 0 to 60 or was more suitable to measure SDB and detect sleep apnea and hypopnea than PM or ODI when the AHI ≤30 (Figure 3).

Monitoring vital signs such as the heart rate (HR), RR, body temperature, urine volume, etc., are important for hospitalized patients, especially in critical care units, and elderly patients at home.^22–24^ Compared to the RR, objective information on HR or body temperature is easily obtained. RR is controlled by not only metabolic but behavioral factors^2,3^ and may be affected not only by a directly attached chest belt but by inspections by medical staff. Therefore, since there is no behavioral control on respiration during sleep, objective RR data can be obtained but problems remain related to the high prevalence of SDB, mostly sleep apnea and hypopnea. Recent data showed that almost 20% of adult males and 10% of menopausal females had moderate (AHI≥15) OSA.^5–7^ Also, prevalence is usually higher in the presence of other diseases.^6^ Therefore, a non-contact and non-constraining RR measuring instrument which can calculate not only the RR while awake but sleep apnea and hypopnea during sleep is desired.^8–11^

Monitoring and analyzing respiratory patterns (normal breathing and abnormal breathing during sleep) using microwave doppler radar is useful and feasible.^8–11^ Indeed, comparisons between radar monitoring and PSG have been done ^12-,15^ but not in clinical settings with PM and their usefulness was not validated. In this study, we showed that a newly developed respiratory monitoring system by MWR could calculate normal breathing at any position correctly and 0≤AHI≤60 as well as a PM or 3% ODI.^17^ The Bland-Altman plot also showed equivalence of our modified radar system to SpO_2_ continuous monitoring when detecting not only AHI from 0 to 60 but <30. It is important for the monitor to detect low AHI levels because generally medical staff or bed partners can visually check for severe SDB (AHI≥30).

A study limitation was the small number of participants although AHIs were equally spread over the ranges examined (0≤AHI≤60). Since this system can detect mild to moderate sleep apnea from Bland-Altman plots from AHI 0 to 30 and mean error data (Figure 3 and Table 2-b), hospital medical staff and caregivers of elderly family members in serious condition could gain information on early changes in patients’ status. Patients’ prognosis worsened when complicated by sleep apnea.^25,26^ Since this is a non-contact system, we cannot determine oxygen levels. However, if we catch SDB early, we can add SpO_2_ monitoring. In addition, for elderly patients at home, continuous RR monitoring with regular SpO_2_ monitoring would be useful because this combination could reveal early or serious changes in pathophysiology. Also, we deleted SDB following 3 min of large movements and one SDB during 6 min. At present, we cannot determine whether this new algorithm is superior to others but the accuracy of RR data including sleep apnea or hypopnea would support its value.

In conclusion, respiratory monitoring by radar with the newly developed algorithms can correctly measure RR while the patient is awake at any position and detect SDB or sleep apnea and hypopnea as well as type 3 portable monitoring or SpO_2_ continuous monitoring. Clinical data accumulated in this clinic indicate that this system would be useful in various medical settings.

## Data Availability

All data produced in the present study are available upon reasonable request to the authors.

## Author Contributions

Conception and design: Chin Kazuo

Analysis and interpretation: Chin Kazuo, Okumura Shigeaki, Endo Daisuke, Nagata Kazuma Collecting the data and drafting the manuscript for important intellectual content: Chin Kazuo, Okumura Shigeaki, Endo Daisuke, Nagata Kazuma, Ito Tatsuya, Murase Kimihiko, Sunadome Hironobu, Hoshi Mamiko, Hiranuma Hisato, Kozu Yutaka, Sato Susumu, Hirai Toyohiro, Gon Yasuhiro, Sakamoto Takuya, Taki Hirofumi, Akahoshi Toshiki

## Acknowledgements

We are grateful to the medical stuffs in Shinjuku Sleep and Respiratory Clinic, KEISHINKINENKAI Medical Corporation. - Tokyo (Japan) for their assistance in performing the study. We are grateful to Tomoko Toki and Mai Henmi for their assistance in preparing the manuscript.

## Conflicts of interest

Kazuo Chin reports funding from Eizai, Aculys Pharma, Magnet, and The Japan Research Foundation for Healthy Aging. He has received consulting fees from Shionogi, Aculys Pharma, and Eli Lilly. Additionally, he reports financial interests related to Philips Japan, Fukuda Denshi, Fukuda Lifetec Tokyo, and ResMed, which have provided funds for the Department of Sleep Medicine and Respiratory Care at his institution.

Shigeaki Okumura reports payment for an oral presentation from Philips Japan at the 22nd CPAP Research meeting. He also holds stock options from MaRI Co., Ltd. through his institution. Kimihiko Murase reports that the department of Advanced Medicine for Respiratory Failure of Kyoto University Graduate School of Medicine is funded by grants from Teijin Pharma to Kyoto University.

Hironobu Sunadome reports funding from Philips Japan, Fukuda Denshi, Fukuda Lifetec Keiji, and ResMed.

Susumu Sato reports funding from Nippon Boehringer Ingelheim. Additionally, Philips Japan, Fukuda Denshi, Fukuda Lifetec Keiji, and ResMed have provided funds to his institution.

Toyohiro Hirai reports funding from Philips Japan, Fukuda Denshi, Fukuda Lifetec Keiji, and ResMed.

Yasuhiro Gon reports that Philips Japan, Fukuda Denshi, Fukuda Lifetec Tokyo, and ResMed Japan have made donations to an endowed department at Nihon University.

Takuya Sakamoto reports royalties from Ohmsha Ltd. and MaRI Co., Ltd. He has received consulting fees from GLORY LTD., Sumitomo Electric Industries Ltd., and Mitsubishi Electric Corporation, with payments made to his institution. He has also received honoraria for lectures and presentations from several organizations, including Science & Technology Co., Ltd., Think&Act, Inc., KEYCOM Corporation, SINSHU UNIVERSITY, The Institute of Electronics, Information and Communication Engineers (IEICE), Sony Semiconductor Solutions Corporation, SENSAIT Council (SAIC), Kyoto University Original Co., Ltd., Japan Human Factors and Ergonomics Society, Kyoto Research Park, Advanced Science, Technology & Management Research Institute of KYOTO, and the Science and Technology in Society (STS) Forum Kyoto Executive Committee. He reports planned, issued, or pending patents with US Provisional Patent Application numbers 63275949 and 63299958.

Hirofumi Taki reports honoraria for lectures from Kyoto University, Advanced Science, Technology & Management Research Institute of KYOTO, Ritsumeikan University, Kyoto Research Park Corp., and the Public Foundation of Kansai Research Institute. He also holds stocks and stock options from MaRI Co., Ltd. through his institution.

No other authors have conflicts of interest to report.

