## Supplemental Files for "Non-contact and non-constraining monitoring of the respiratory rate including sleep disordered breathing using ultra-wideband radar"

**Supplemental Data**

**Figure legends**

Figure S1.

Respiratory patterns according to the ratio of two (normal breath and apnea or hypopnea) Gaussian distributions in the Expectation-Maximization algorithm

(a) Measurements by ribcage and abdominal respiratory inductance plethysmographs, (b) Measurements by the flow sensor and (c) Displacements of movements by the radar.

When the ratio of two (normal breaths and apnea or hypopnea) Gaussian distributions in the Expectation-Maximization algorithm was 0.93, there was no apnea or hypopnea. When it was 0.32, there was overt apnea or hypopnea and we made the threshold 0.70.

EM: Expectation-Maximization

Figure S2.

Comparison of respiration rates (RR) of participants with and without a blanket.

Orange solid line with circles shows the manually counted RR. Gray solid line with cross points shows results of radar measurements. Methods for calculation of RR by of radar or manually were the same as in Fig 1-d.

Figure S3.

Scatter plots between AHI and REI by radar or PM and the 3% ODI.

Each correlation was significant ($p<0.001$).

AHI: apnea hypopnea index, REI: respiratory event index, PM: portable monitoring, ODI: oxygen desaturation index, MWR: millimeter wave radar.

**Figure S1.**

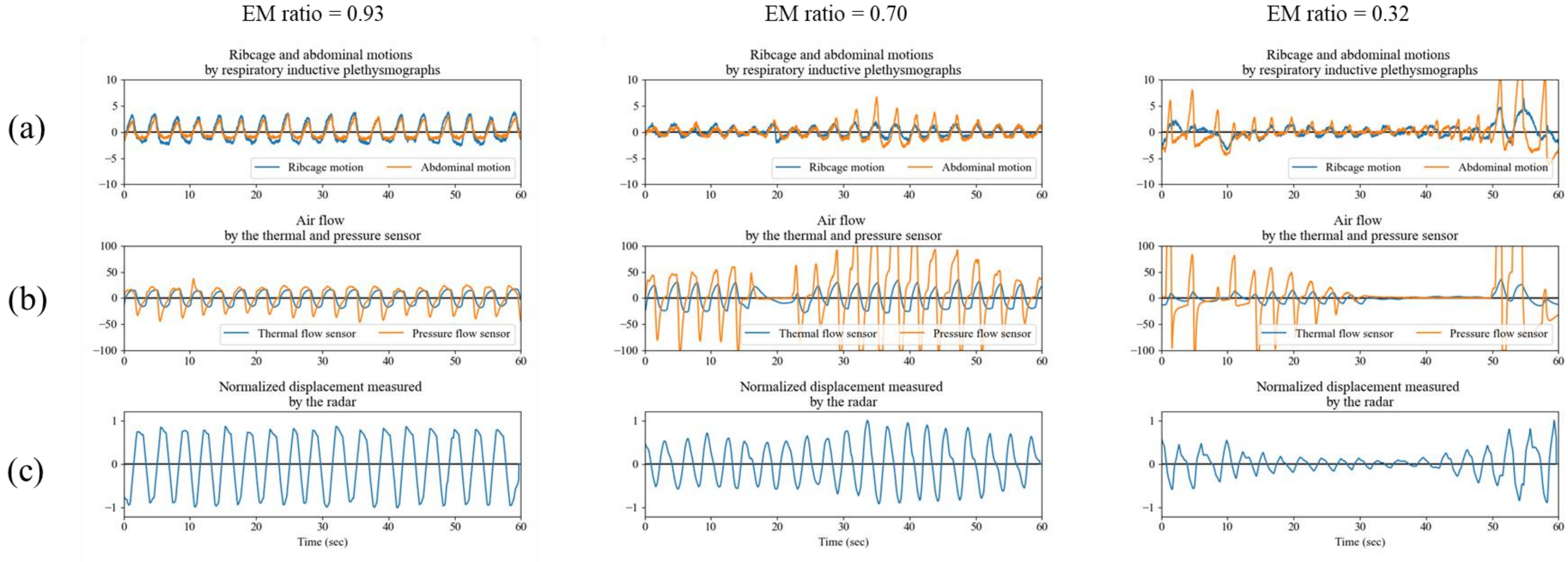
EM ratio = 0.93
EM ratio = 0.70
EM ratio = 0.32
(a)
(b)
(c)
Ribcage and abdominal motions
by respiratory inductive plethysmographs
Ribcage motion
Abdominal motion
Air flow
by the thermal and pressure sensor
Thermal flow sensor
Pressure flow sensor
Normalized displacement measured
by the radar
Time (sec)

**Figure S2.**

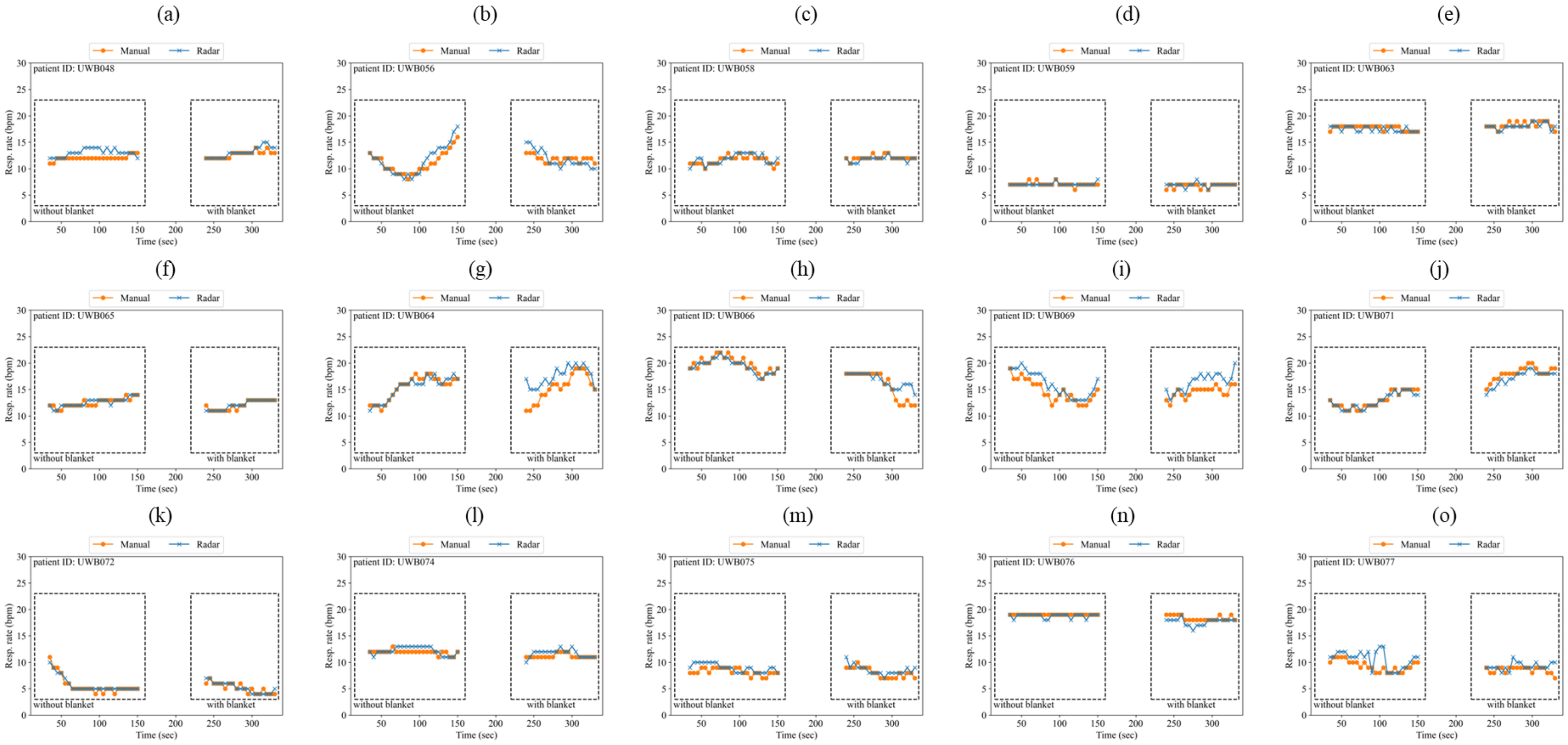
(a)
Manual
Radar
patient ID: UWB048
without blanket
with blanket
Resp. rate (bpm)
Time (sec)
(b)
patient ID: UWB056
(c)
patient ID: UWB058
(d)
patient ID: UWB059
(e)
patient ID: UWB063
(f)
patient ID: UWB065
(g)
patient ID: UWB064
(h)
patient ID: UWB066
(i)
patient ID: UWB069
(j)
patient ID: UWB071
(k)
patient ID: UWB072
(l)
patient ID: UWB074
(m)
patient ID: UWB075
(n)
patient ID: UWB076
(o)
patient ID: UWB077

**Figure S3.**

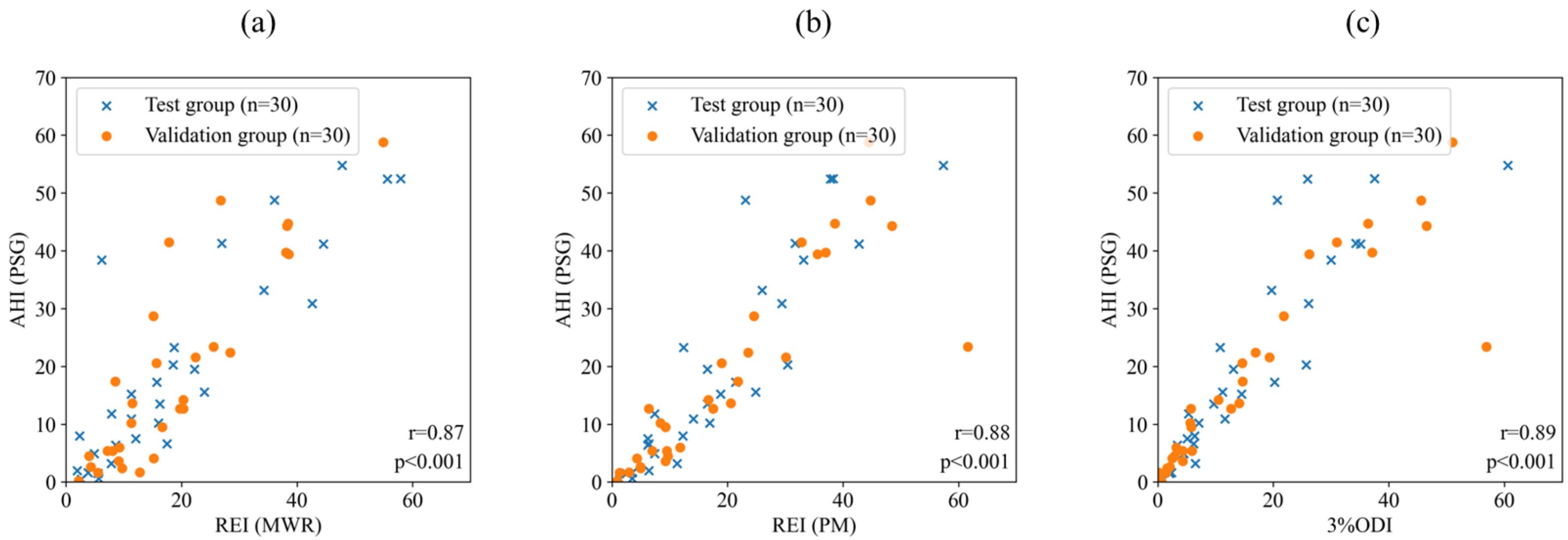
(a)
(b)
(c)
Test group (n=30)
Validation group (n=30)
AHI (PSG)
REI (MWR)
REI (PM)
3%ODI
r=0.87
p<0.001
r=0.88
p<0.001
r=0.89
p<0.001
0
10
20
30
40
50
60
70
